# Prenatal and postnatal environmental influences on child growth trajectories across a rural-urban gradient: an analysis of the ECoMiD longitudinal birth cohort

**DOI:** 10.1101/2025.11.10.25339957

**Authors:** Caitlin Hemlock, Gwenyth O. Lee, Benjamin F. Arnold, Jesse D. Contreras, Molly K. Miller-Petrie, Gabriel Trueba, Andrea Anchundia, Nancy Castro, April M. Ballard, Joseph N.S. Eisenberg, Karen Levy, Karen Levy, Joseph N.S. Eisenberg, Gwenyth O. Lee, Gabriel Trueba, Benjamin F. Arnold, Kostas T. Konstantinidis, William Cevallos, Field data collection subgroup, Adriana Lupero, Mauricio Ayoví, Molly K. Miller-Petrie, Data management subgroup, Jesse Contreras, Jessica Uruchima, Nutrition Subgroup, Nancy Castro, Andrea Anchundia the ECoMiD Authorship Group

## Abstract

**Background:** Early childhood growth, a key indicator of child well-being, is shaped by both the prenatal and postnatal environment. To understand where and when environmental factors influence growth most, we analyzed size at birth, postnatal growth trajectories, and associations with environmental factors across a rural-urban gradient in the ECoMiD longitudinal birth cohort.

**Methods and Findings:** Between 2021-2022, pregnant women were enrolled from four sites in northwest coastal Ecuador (urban city, intermediate town, rural villages connected by road, rural villages connected by river) and children were visited shortly after birth and quarterly thereafter for 24 months. We selected exposure variables using UNICEF’s Conceptual Framework on Maternal and Child Malnutrition, including demographics, maternal characteristics, household infrastructure, and environmental factors. We used multivariable linear models and generalized estimating equations to analyze associations between exposures and length-for-gestational-age Z-score (LGAZ) and postnatal change in 3-month length-for-age Z-score (3-month ΔLAZ) and explored effect measure modification by site and child age (0-6 versus 7-24 months). Among 401 children, LGAZ was higher among children born to taller mothers, in households with higher food security, and in the rural river site. However, the postnatal growth of children in rural river sites faltered most rapidly, particularly after six months, resulting in 51% of children stunted (LAZ < -2 SD) at 24 months, compared to urban (8%), intermediate (14%), rural road (5%). Over the 0-24 month postnatal period, 3-month ΔLAZ was negatively associated with children who were male, from poorer households, had shorter mothers, and had greater exposure to animals and animal feces. Other associations, including with season, hygiene characteristics, and maternal demographics, varied by age and site.

**Conclusions:** These findings show that environmental conditions influenced postnatal linear growth more strongly, compared to prenatal growth, which was associated with maternal anthropometry and nutrition. Community context strongly influenced growth only after age six months and modified associations with environmental conditions, suggesting that interventions targeting child growth faltering after six months may be important for certain populations.

## BACKGROUND

Despite progress on improving growth outcomes globally, 150 million children under five still failed to reach their full growth potential in 2024 (1). The first 1000 days, or the prenatal and postnatal period up to two years of age, is recognized as a critical window for growth in early childhood (2). Growth faltering during this period is difficult to reverse, particularly in low-resource settings (3), and is associated with poor long-term health and economic outcomes (4), and can persist across generations (5).

The causes of poor child growth are multifactorial and include prenatal and parental characteristics, birth characteristics, the household environment, and the broader social, economic, and geographical context of the community (6,7). A commonly used framework to understand relationships between causes of poor child growth is the UNICEF Conceptual Framework on Maternal and Child Nutrition, which illustrates how broader structural factors, which are most difficult to intervene on, operate through household and caregivers factors, and ultimately proximal exposures related to nutrition and care (6). Recent studies exploring these factors found that maternal anthropometry and child birth size accounted for the largest effects on attained growth, while household and environmental variables, such as socioeconomic status and water, sanitation and hygiene characteristics, have smaller effects (7–9). However, even if maternal factors have the largest effects on attained growth, preventing growth faltering for subsequent generations may still require interventions targeting the early childhood period, in addition to the prenatal period. Understanding not only which exposures, but when and where they are most influential on growth faltering is important to inform the targeting of interventions during this critical window of growth. Comparing growth trajectory differences between settings within a country can help control for political, social, and cultural differences that may explain variation in structural drivers when examining across countries, as other studies in multiple settings have done.

Our objective was to characterize child growth trajectories from birth to two years in the ECoMiD (‘Enteropatógenos, Crecimiento, Microbioma, y Diarrea’) birth cohort (10). The ECoMiD cohort is unique as it comprises of four sites in a resource-poor area of northwest Ecuador with varying levels of urbanicity, offering the ability to examine the impact of varying environmental and geographic conditions on longitudinal growth. We analyzed associations between child growth trajectories and environmental, behavioral, and demographic characteristics at the community and household level. To understand when characteristics had the strongest effect, we assessed differences across specific developmental periods. We also assessed if there were differences in the influence of environmental exposures on growth trajectories by urbanicity.

## METHODS

### Study design

ECoMiD was a prospective, longitudinal birth cohort study conducted from 2019-2024. We recruited pregnant women in the 3rd trimester and collected data and biological samples from them (*in utero* when recruited) and their child and environmental samples from the household. Samples were collected during pregnancy at enrollment, one week after birth, and every three months until 24 months. Enrollment from ten communities along a rural-urban gradient ensured that children experienced a range of environmental conditions. Communities included Santo Domingo, Zancudo, Colón de Onzole, and San Francisco (rural villages only accessible by river or with limited car accessibility, and populations of ∼200–700 each; referred to as ‘rural river’ sites); Timbiré, Selva Alegre, Colón Eloy, and Maldonado (rural villages accessible by road near Borbón, populations ∼500-1000 each; referred to as ‘rural road’ sites); the town of Borbón, the commercial center of Borbón parish (population ∼5,000; referred to as the ‘intermediate’ site); and the city of Esmeraldas, capital of Esmeraldas province (population ∼162,000; referred to as the ‘urban’ site) (Figure 1).

**Figure 1.**
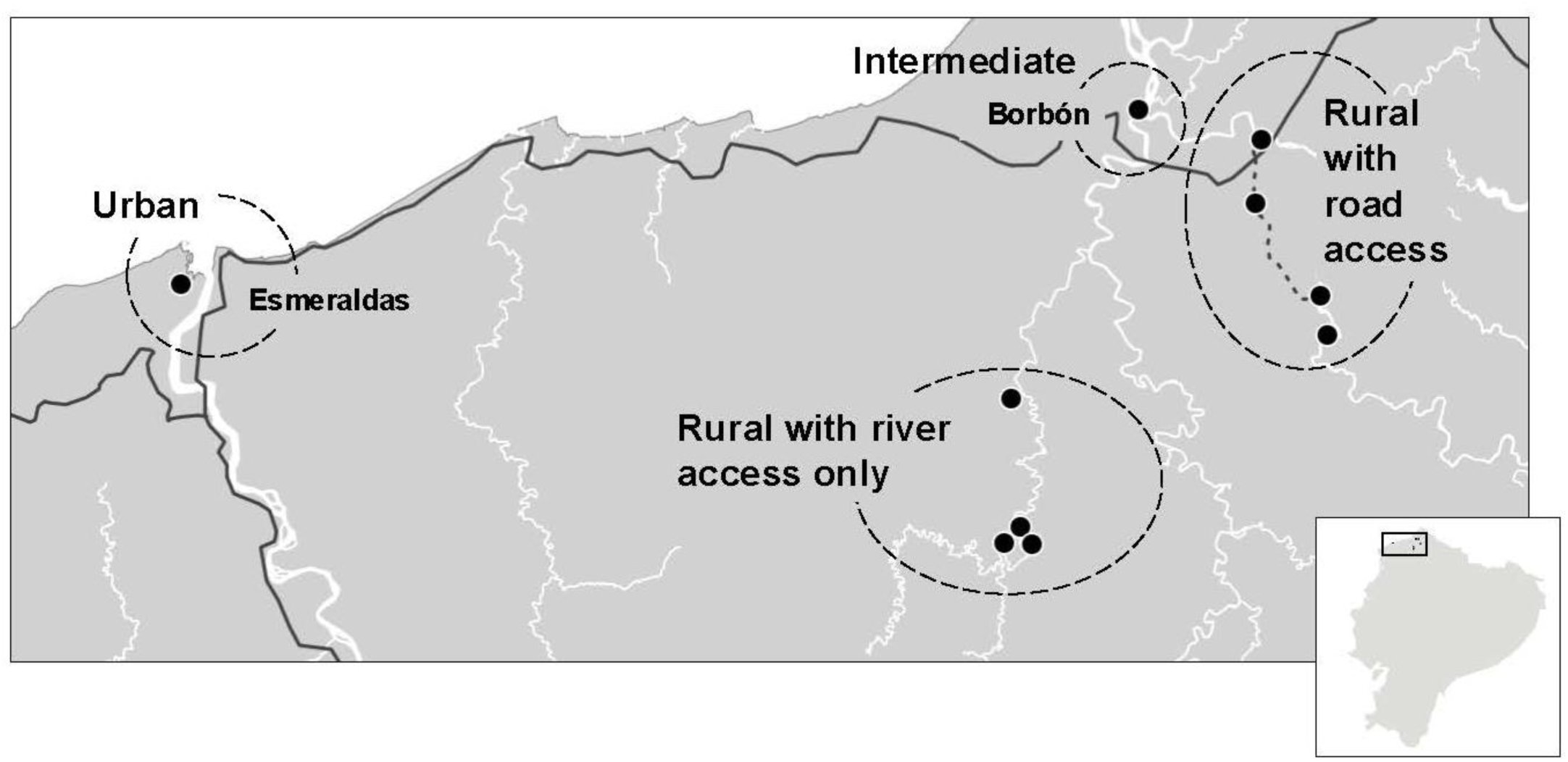
Map of study sites in the ECoMiD cohort in northwestern coastal Ecuador.

### Participants

Detailed information about the ECoMiD cohort, including recruitment and enrollment, have been published previously (10). Recruitment began in 2019, however enrollment and in-person data collection were paused in 2020 due to the COVID19 pandemic. For this analysis, we focused on children born to term after the pause in enrollment, starting in February 2021. We also excluded mothers who self-identified as indigenous, as their growth trajectories and household conditions were substantially different compared to the rest of the cohort.

### Ethical considerations

The study protocol was approved by the institutional review boards of the Ecuadorian Ministry of Health (MSPCURI000253-4), the Universidad San Francisco de Quito (USFQ; 2018–022M), the University of Washington (UW; STUDY00014270), and Emory University (IRB00101202).

### Data collection

The study conducted one pregnancy visit shortly after enrollment (∼37 weeks gestation), where data on household demographics, including maternal age, self-reported maternal ethnicity, highest level of maternal education, household assets, and water, sanitation and hygiene infrastructure, were collected. At the initial visit during pregnancy and follow-up visits conducted every six months, enumerators asked about food security (measured by the Household Food Insecurity Access Scale (11)) and maternal exposure to animals around the household, as well as child exposure to animals starting at six months. Enumerators also collected environmental samples, including drinking water and hand rinses from mothers and the target child starting at six months, which were tested for *E. coli* using Colilert and Petrifilm (methods described previously (10,12)). Enumerators also performed spot checks of the household to assess the presence of feces, garbage, and other variables associated with hygiene. Maternal height and weight was measured six months postpartum, however maternal body mass index (BMI) was not included in the analysis because of high levels of missingness for weight. The study also conducted weekly surveillance visits, where enumerators asked about breastfeeding habits and if the child consumed breastmilk, formula, liquids other than breastmilk, and solid food for each day in the past seven days. All data were collected on tablets using the Open Data Kit platform (https://opendatakit.org).

To obtain birth anthropometry, enumerators requested to see a physical copy of the child’s official birth card at the 1-week visit and entered birth length, birth weight, and gestational age into the ODK platform. Photos were also captured in the ODK platform for later quality control via optical character recognition (OCR). If estimated gestational age was not available on the birth card, we used maternal-reported gestational age at enrollment and date of birth to calculate gestational age at birth. Postnatally, child recumbent length and weight was measured in duplicate, and if the margin of error was greater than 0.9cm, a third measurement was taken. If biologically implausible values were identified, enumerators returned to the household to remeasure.

### Conceptual Framework

We organized the environmental characteristics for the available data according to the UNICEF Conceptual Framework on Maternal and Child Nutrition (Figure 2), which we also theorized could function as directed acyclic graph for the connections between the characteristics and child growth. Characteristics were mostly collected at the household-level, however we considered some as proxies for broader determinants. Within the framework, determinants are grouped into three categories: *Enabling Determinants*, *Underlying Determinants,* and *Immediate Determinants*.

**Figure 2.**
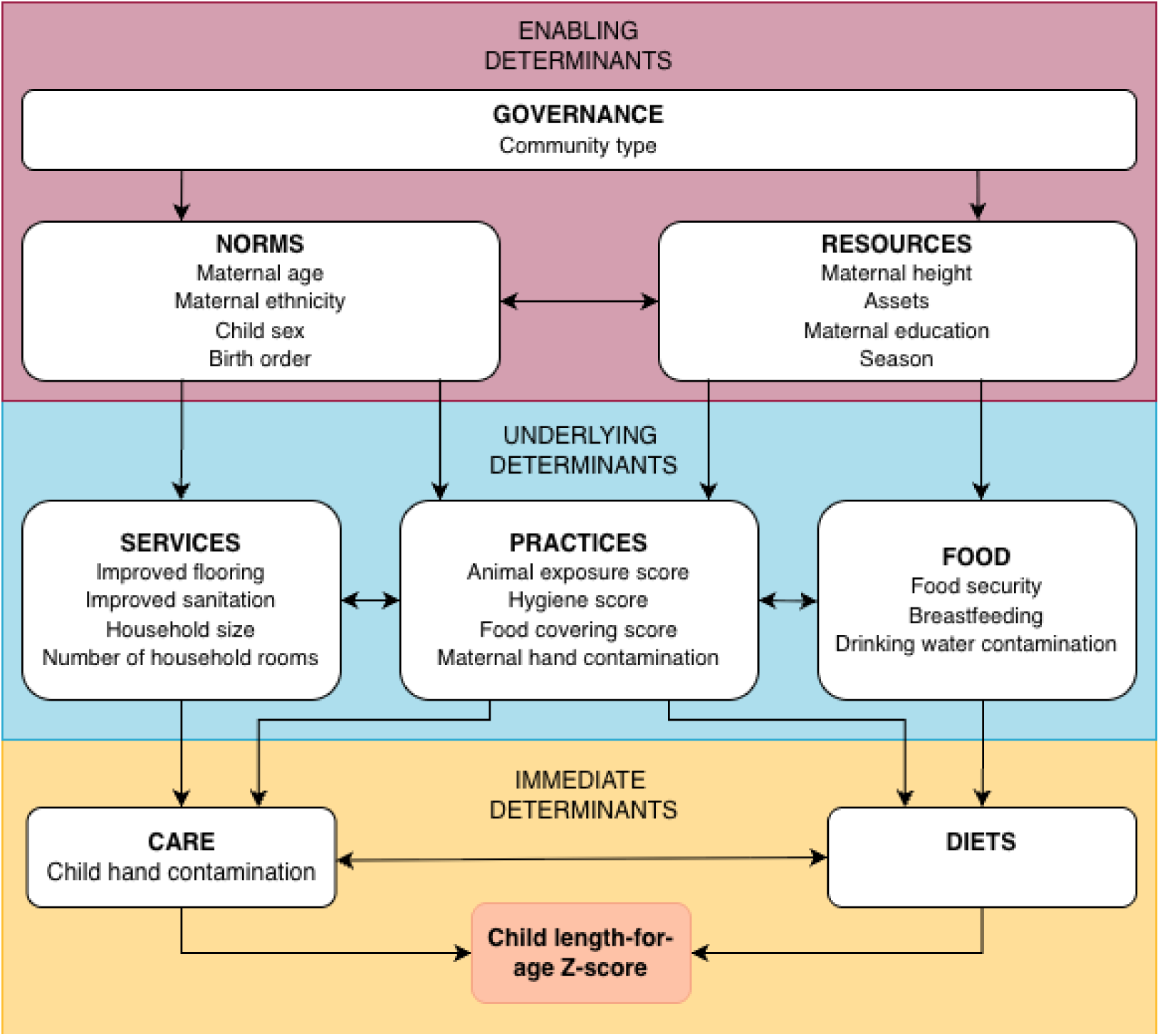
Community and household characteristics assessed in ECoMiD mapped onto the UNICEF Conceptual Framework on Maternal and Child Nutrition (6).

*Enabling Determinants* were variables hypothesized to be proxies of *Governance*, *Resources*, and *Norms* of the study area. <u>Community type</u> was considered a proxy of *Governance* and was divided into the four community types from the cohort. For *Resources*, we included <u>maternal heigh</u>t, potentially a representation of generational availability of resources resulting in stunting during the mother’s early life; <u>household wealth</u> (defined by an asset score from first component extracted from a multiple correspondence analysis of 11 assets (13)); <u>maternal</u> <u>education during pregnancy</u> (categorized into “Primary or less” [six years or less of education], “Lower secondary” [7-9 years], “Upper secondary” [10-12 years], “Post-secondary or greater” [more than 12]); and <u>season</u> (categorized into early rainy [January-March], late rainy [April-June], early dry [July-September], and late dry [May-December]). We divided season into early and late rainy and dry to capture broad variation in factors including risk of enteropathogen infection, access to employment, resources, and food. For *Norms*, we considered physical attributes that are indicators of social norms, including <u>child sex</u>, potentially representing socio-cultural gender norms in addition to biological differences in risk; <u>birth order</u> (categorized into first, second, and third or more); <u>maternal age during pregnancy;</u> and self-reported <u>maternal ethnicity</u> (dichotomized to Afro-Ecuadorian and other, typically mestizo or Manabí).

*Underlying Determinants* included variables related to *Food*, *Practices*, and *Services*. For *Food*, we included <u>food</u> <u>security</u> as a binary variable of moderately/severely insecure vs. mildly insecure/secure; and whether or not the child was <u>exclusively breastfed</u> during the first six months. Using daily data on infant feeding, days were categorized as exclusive (infant receiving breastmilk with no other food except syrup medicine), predominant (infant receiving only breastmilk for nutrition and water or water-based drinks like herbal tea), or partial breastfeeding (infant receiving non-human milk feeds such as animal milk, formula milk, or other food) (14). The proportion of days with available data where the infant was exclusively breastfed was binarized into ≤50% (0) or >50% of days (1). We also evaluated <u>drinking water contamination</u>, which was an average of fecal contamination measured as *E. coli* in drinking water samples collected during the pregnancy visit and at 6, 12, and 18 months, with three measurements taken per timepoint (details in SI). For *Practices*, we included an ordinal <u>household</u> <u>hygiene score</u> collected during pregnancy; an ordinal <u>food hygiene score</u> collected during pregnancy; and a continuous <u>animal exposure score</u> collected during pregnancy and at 6, 12, and 18 months and scaled to range from 0-10. Methods for these three variables have been published previously (12). We also included <u>maternal</u> <u>hand contamination</u>, constructed similarly to drinking water quality (details in SI). For *Services*, we included <u>number of household members</u>; <u>number of rooms</u> in the household; a binary variable for <u>improved flooring</u> (earth/sand/palm/bamboo flooring versus finished flooring); and a binary variable for <u>improved sanitation</u> (Basic/Limited sanitation versus Unimproved/Open sanitation according to the Joint Monitoring Program service ladder (15)).

*Immediate Determinants* are related to *Diets* and *Care*. Since we were interested in environmental variables for this analysis, we included one environmental exposure of interest, a semi-continuous score of <u>child hand</u> <u>contamination</u> averaged over measurements at 6, 12, and 18 months with three measurements taken per timepoint (details available in SI). We included child hand contamination here as we hypothesized it would be downstream of maternal hand contamination and represent a dimension of *Care*.

Because exposure variables were collected at different intervals by design, any variables where the interval between data collection was greater than three months were imputed by carrying data forward (Figure S1). This was only done for data missing by design (i.e. if a variable was missing at a timepoint where data was supposed to be collected, we did not carry forward the value from the previous non-missing visit). Two exceptions include animal score, where we imputed maternal animal exposure score from the six-month visit to represent child exposure at 1 week and 3 months, and food security, where we carried data collected at the 12-month visit through to the 24-month visit as data collection stopped for this variable at 12 months.

### Outcomes

Our two outcomes of interest were attained length at birth and change in linear growth postnatally. For <u>attained</u> <u>length at birth</u>, we used birth length and gestational age to calculate length-for-gestational-age Z-score (LGAZ) using INTERGROWTH standards (16). For <u>change in growth postnatally</u>, we used the R package *zscorer* to calculate length-for-age (LAZ) based on WHO 2006 growth standards and excluded extreme Z-scores above 6 SD and below -6 SD (17). We used the package *growthcleanr*, which uses exponentially-weighted moving averages to remove implausible measurements and inconsistencies across individual trajectories of height and weight (18). We calculated change in LAZ for each three-month postnatal interval (3-month ΔLAZ) where Z-score was available at the beginning and end of the interval.

### Statistical Analysis

We ran separate models for LGAZ and 3-month ΔLAZ. We used generalized linear models to obtain the mean difference in LGAZ for a given exposure, adjusting for covariates as specified below. For 3-month ΔLAZ, we used generalized estimating equations with an exchangeable working correlation structure to obtain mean differences while controlling for repeated measures within the same child and included robust standard errors clustered at the child-level.

Our covariate adjustment strategy used the variables mapped onto the UNICEF framework as a modified directed acyclic graph (Figure 2). For each exposure, we ran a model that included all exposures at the same level of the UNICEF framework and all upstream variables (for example, to obtain the parameter of interest for *Underlying Determinant* exposures, we included all variables at the *Underlying Determinant* level and all *Enabling Determinants*). At the *Underlying Determinant* level, we used a separate model for environmental contamination variables as we hypothesized this would be downstream of most variables included at that level of the framework. Thus, estimates for environmental contamination variables are adjusted for all other *Underlying* Determinants, but not vice versa. We did this for both outcomes of interest (LGAZ and 3-month ΔLAZ). We adjusted for time fixed effects (year) in all models and age (months at the beginning of the growth interval modeled with natural cubic splines with knots at 6, 12, and 18 months) in the models for 3-month ΔLAZ. We also adjusted for LAZ at the beginning of the interval to account for regression to the mean and days between measurements to account for measurement differences in 3-month Δ LAZ models. We used complete case analysis and assumed that any missingness or loss to follow-up was not associated with unobserved variables (missing at random).

To understand whether associations differ by urbanicity and child developmental period, we refitted regression models for each exposure, including an interaction term with these effect modifiers. For urbanicity, we used community type (urban, intermediate, rural road, and rural river) and examined effect modification for both LGAZ and 3-month ΔLAZ. For child developmental period, we considered 6 months as a cut-point, given the differences with nutrition and environmental interactions across the two developmental periods. We created an indicator of whether the beginning of the 3-month change period was 0-6 months or >6 months and looked at effect modification for 3-month ΔLAZ. We used Wald tests to test the hypothesis of overall interaction (p < 0.05 for significant interaction) and predicted marginal effects by developmental period and community type.

We performed all statistical analyses in R version 7.2. Code is available on GitHub (insert URL after acceptance).

## RESULTS

### Study sample and characteristics

We enrolled 404 pregnant mothers in the ECoMiD cohort after the COVID enrollment pause, from February 2021 to December 2022. Two mothers who self-identified as indigenous and their children were excluded, as well as one infant born preterm (Figure 3). Among the 401 included, five mothers (1.2%) did not have live births or withdrew prior to delivery. Among the remainder, 74 (18.7%) were lost to follow-up over the course of the 24-month period, with attrition ranging from 4-15 children per time point. Additional missing anthropometry data resulted from mothers not being available for household visits, temporary migration from the study area, or invalid or biologically implausible LAZ values (Figure 3).

**Figure 3.**
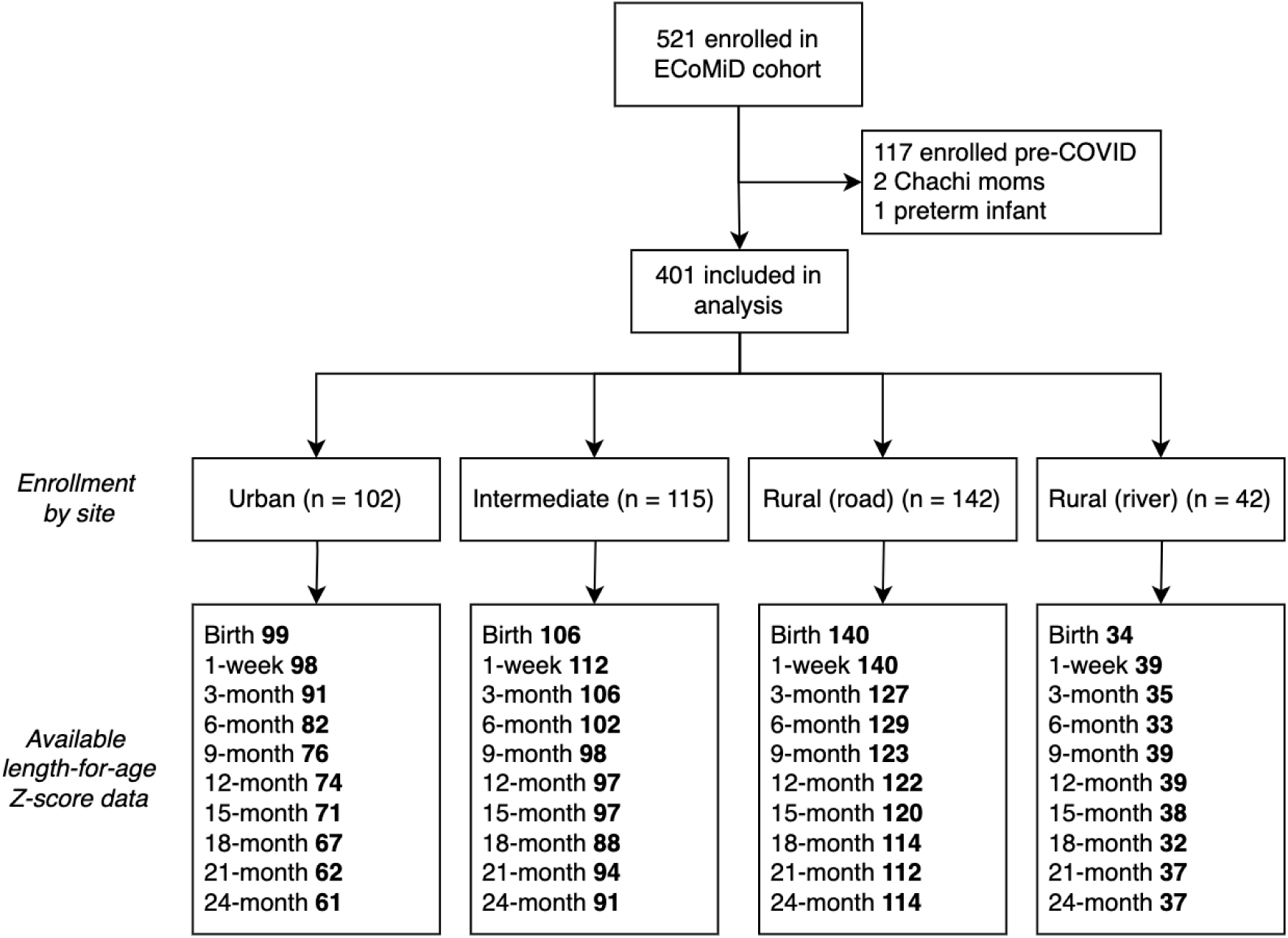
Study flow chart and available linear growth data in the ECoMiD cohort. Note: Site enrollment numbers are pregnant women enrolled. Numbers for Birth include those born alive with length and gestational age data available on birthcards and biologically plausible length-for-gestational-age Z-score (-6 < SD < 6). Data for each subsequent timepoint are children with available and biologically plausible length-for-age-Z-score data.

Mothers in rural areas were generally older and less educated, had higher parity, and were more likely to identify as Afro-Ecuadorian (Table S1). Households in rural areas were less likely to be in a higher wealth quartile, while households in the rural river site were less likely to have improved flooring or an improved toilet. In contrast, hygiene scores, household size, and maternal height were relatively consistent across sites. Half of children were exclusively breastfed for more than 50% of days in the first six months, which was consistent across sites except rural river, where three quarters of children were exclusively breastfed. Food security during pregnancy was highest in the rural road site and average food security across sites was consistent through visits (Table S2), while average exposure to domestic animals increased with age.

### Growth curves

Birth size and postnatal growth trajectories varied across settings. Children in rural river sites were born the largest (Table S1) but their LAZ faltered more rapidly than in the other sites, particularly after six months (Figure 4). The mean ΔLAZ in the rural river site was -0.24 (0.89 SD) from 0 to 24 months, compared to -0.13 (0.81 SD) across all settings (Table S1). These losses in linear growth accumulated, resulting in the prevalence of stunting or severe stunting at 24 months of 51% in the rural river site, compared to 15% in the intermediate site, 8% in the urban site, and 5% in the rural road site. Variation by site and by age in LAZ and stunting accumulation over time is shown in Figure 4.

**Figure 4.**
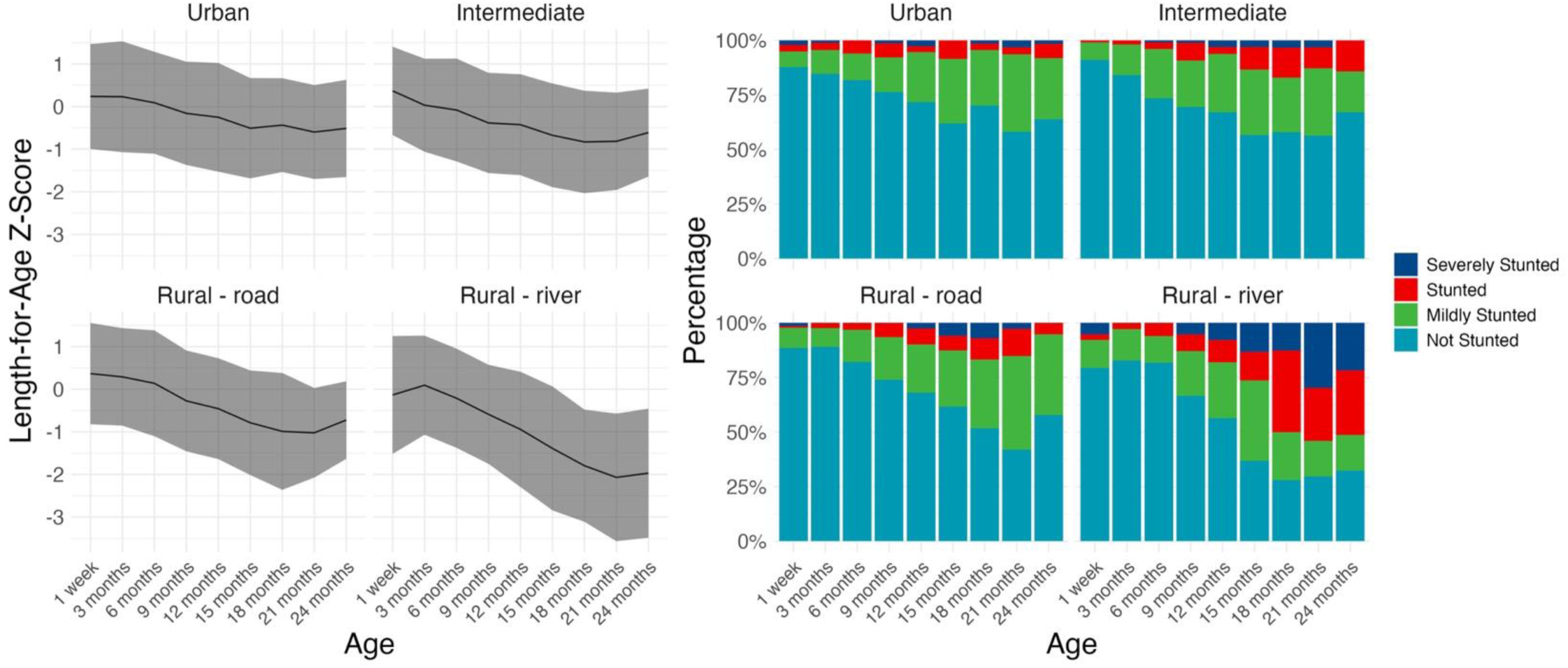
Mean length-for-age Z-score (LAZ) and percentage of children mildly, moderately, and severely stunted, by timepoint and community type. Note: Ribbons in line graph are standard deviation by visit. Severely stunted = <-3 SD below age- and sex-specific reference median; stunted = <-2 SD below median; mildly stunted = <-1 SD below median; not stunted = ≥-1 SD below median.

### Associations with birth size

Among the 20 exposures we assessed (Figure 2), only maternal height and food insecurity were significantly associated with LGAZ (Table 1). Taller maternal stature was associated with higher LGAZ, with an increase of 0.18 SD per 5 cm maternal height (95% CI: 0.06, 0.25), while children born in households experiencing food insecurity during pregnancy were associated with a 0.32 SD lower LGAZ (95% CI: -0.62, -0.01).Children born in rural road and rural river sites were largest, however neither were statistically significantly different from children born in the urban site (rural-road mean difference [MD]: 0.31 SD, 95% CI: -0.002, 0.62; rural-river MD: 0.34 SD, 95% CI: -0.13, 0.81).

**Table 1.**
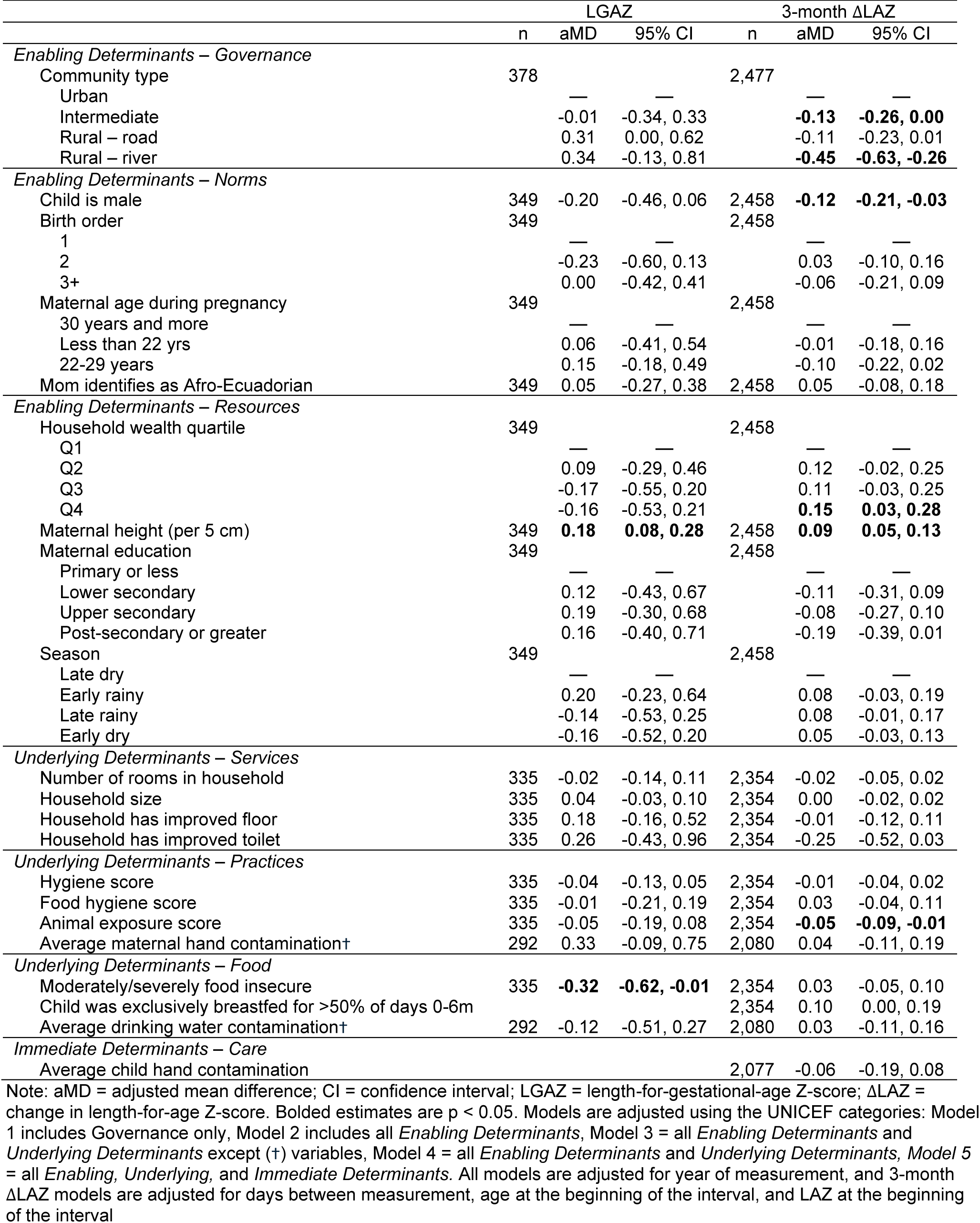
Associations between community, demographic, and environmental exposures with child anthropometry at birth and postnatal growth trajectories, grouped by category in the UNICEF Conceptual Framework on Maternal and Child Nutrition (6) (See also Figure 2).

The associations between maternal height and LGAZ and food security and LGAZ did not significantly vary by community type (p = 0.28, 0.98, Table S4). The only exposure variable that had no overall association with LGAZ, but significant interaction with community type, was number of rooms in the household (p = 0.04), where a larger household was more beneficial for birth size for households in rural-river communities (0.31 LGAZ MD; 95% CI: 0.04, 0.58).

### Associations with postnatal growth trajectories

Children residing in the intermediate, rural road, and rural river communities had larger decreases in 3-month ΔLAZ during the postnatal period than children residing in the urban site (Table 1), and this was more pronounced for children residing in the rural river communities during the 7–24-month period (Table S5). Male children had larger decreases than female children (-0.12 ΔLAZ, 95% CI: -0.21, -0.03) and children born in the wealthiest households had a growth advantage compared to those born in the lowest quartile of wealth (Q4 vs. Q1: 0.15 ΔLAZ, 95% CI: 0.03, 0.28) (Table 1), with no differences across child age or community type (Table S5, Table S6). Maternal height was positively associated with ΔLAZ across child developmental periods and community types (Table 1).

The only household environmental variable associated with ΔLAZ across the two-year period and communities was animal exposure, with a per point increase in the animal exposure score associated with -0.05 ΔLAZ (95% CI: -0.09, -0.01). The interquartile range for the animal exposure score was 1.4 points (range 0-10), meaning the 3-month ΔLAZ comparing the 25^th^ percentile to the 75^th^ percentile of the animal exposure score distribution in the population was -0.07 ΔLAZ (95% CI: -0.12, -0.02). Although interaction with community type was not significant, the magnitude of association was larger in the urban site compared to the other communities (Table S6).

Several environmental exposures had no overall association with ΔLAZ but varying associations across developmental periods and community type (Figure 5, Table S5, Table S6). Seasonal effects on ΔLAZ were more pronounced during the 7–24-month compared to the 0–6-month, with measurements in the early rainy and early dry period associated with better ΔLAZ compared to the late dry period (early rainy MD: 0.13, 95% CI: 0.01, 0.25, early dry MD: 0.1, 95% CI: 0.01, 0.2) (Figure 5, Table S5). Negative associations with ΔLAZ in the late dry period were driven by children in the rural-road communities, while the children in the rural-river communities had worse ΔLAZ during the late rainy season (-0.27, 95% CI: -0.53, -0.01), and marginally during the early rainy season; no associations were observed with season in the urban or intermediate communities. Child hand contamination was associated with greater decreases in ΔLAZ during the 0-6 month period only (-0.18 ΔLAZ, 95% CI: -0.35, -0.05) (Table S5). Food hygiene was positively associated with ΔLAZ in the rural river site (0.19 ΔLAZ per point, 95% CI: 0.02, 0.35), but differing from our hypotheses, better hygiene scores were negatively associated with ΔLAZ in the rural river communities (-0.27 ΔLAZ, 95% CI: -0.45, -0.09) and food insecurity was positively associated with ΔLAZ in the rural river communities (0.4 ΔLAZ, 95% CI: 0.2, 0.6), while neither variable was associated with ΔLAZ in other community types.

**Figure 5.**
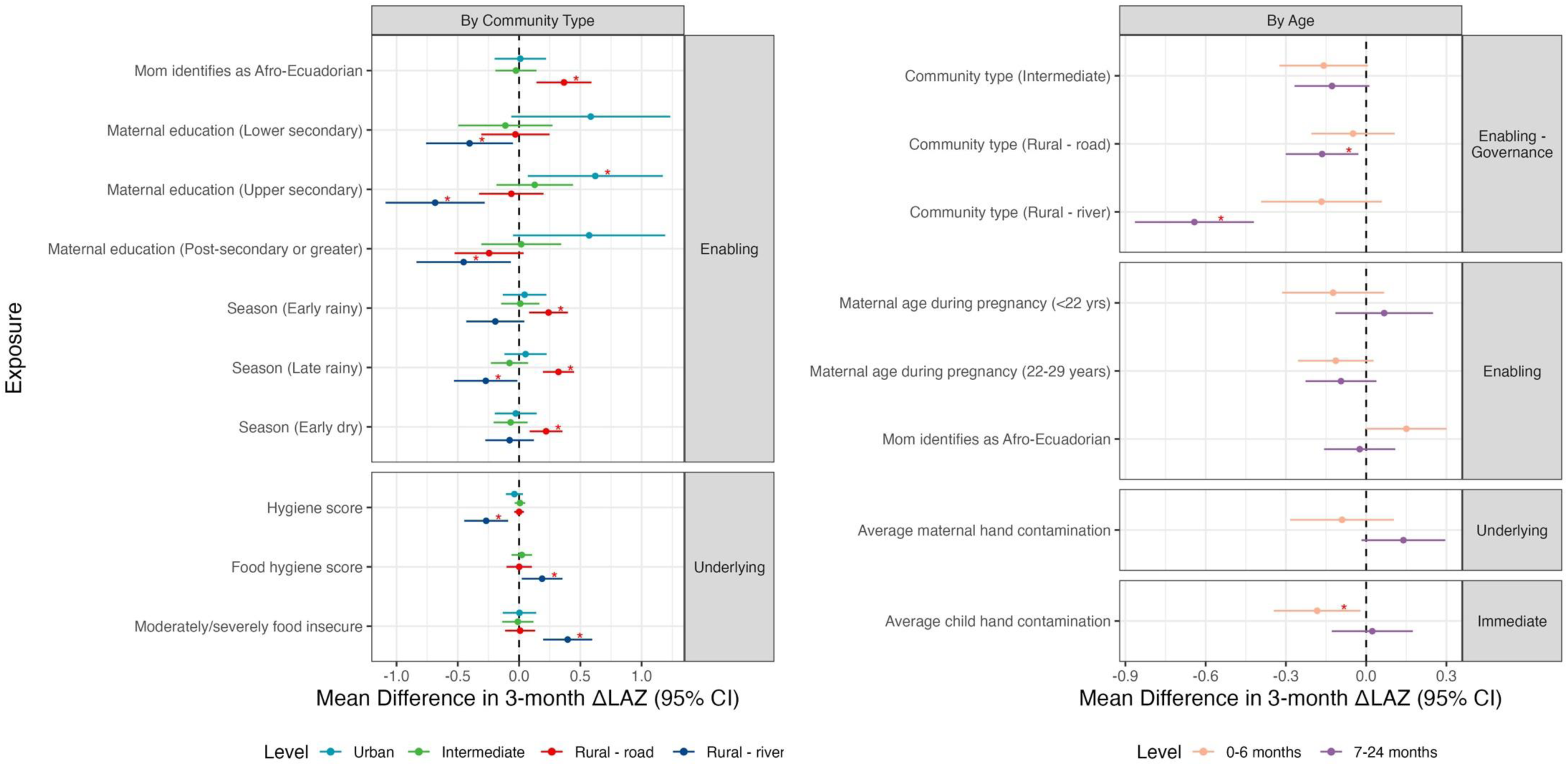
Effect measure modification of associations between environmental, behavioral, and demographic variables and postnatal linear growth trajectories, by age and community type. Note: Estimates displayed are those with significant interaction (F-test p-value < 0.05). Red asterisks indicate if the stratified estimate was significantly different from the null (p-value < 0.05). Reference values for categorical variables are Urban (Community type), 1^st^ born (Birth order), 30 years and older (Maternal age during pregnancy), Q1 (Household wealth quartile), Primary or None (Maternal education), Late dry (Season).

Maternal characteristics were differentially associated with ΔLAZ across communities as well (Figure 5). During the 7–24-month period, children born to the youngest mothers in the cohort had better ΔLAZ compared to the oldest mothers during the same period (Table S5). However, children of the youngest mothers had worse ΔLAZ in the intermediate and rural river communities (Table S6). Higher levels of maternal education were more positively associated with ΔLAZ in the urban sites, while higher education was associated with worse ΔLAZ in the rural-river communities and marginally worse ΔLAZ in the rural-road communities. Children of Afro-Ecuadorian mothers were associated with better ΔLAZ in the 0–6-month, and this association was driven by children in the rural-road communities (0.37 ΔLAZ, 95% CI: 0.14, 0.59).

While exclusive breastfeeding in the first 6 months was only marginally associated with ΔLAZ over the 0-24 month period (0.10, 95% CI: 0.0, 0.19), children in rural-road and rural-river communities benefited most (rural-road MD: 0.2, 95% CI: 0.06, 0.35; rural-river MD: 0.3, 95% CI: -0.07, 0.68), though interaction was not significant (p = 0.12) (Table S6).

## DISCUSSION

In this study, we leveraged the unique design of the ECoMiD birth cohort across a rural-urban gradient in northwest Ecuador to examine child growth trajectories, associations with environmental risk factors, and differences by child age and geographic setting. In this Latin American cohort, we found that growth faltering appeared to be driven by postnatal factors as much, if not more than prenatal factors, suggesting that the relative importance of growth restriction in each period may be context specific. The range of postnatal trajectories we observed across study sites are representative of the range observed across countries with varied structural and cultural factors (3), showing how growth patterns seen at a global scale can present locally. Additionally, associations with household and environmental variables and growth faltering differed by child age and study site within this cohort, showing how growth faltering can develop from different pathways depending on the context. This highlights the importance of targeting interventions that are environmentally focused not only by developmental stage, but geographic location, during the critical period for growth.

### Enabling Determinants

Community type across the rural-urban gradient, used to characterize the broadest exposure in the UNICEF framework (Governance), was strongly associated with both length at birth and postnatal growth trajectories. Children in the intermediate site were born similar size to urban children but experienced more postnatal growth faltering. Children in the rural communities were born larger, but postnatally, children in communities accessible by road had growth trajectories similar to intermediate children, while children in communities accessible by river experienced more severe postnatal growth faltering, resulting in half of children stunted. This disparity in growth trajectories across the rural-urban gradient adds to the body of research showing that children from rural areas have higher rates of stunting than urban areas (19). A study of 23 countries in sub-Saharan Africa found that harmful effects of remoteness are mediated by education, wealth, social services, and infrastructure (20). When we adjusted our community type model for downstream enabling factors at the household level, such as wealth and education, the associations were slightly attenuated but mostly unchanged (data not presented), leading us to hypothesize that there are likely other community-level drivers explaining the variation in growth trajectories by community type.

In addition to differences in postnatal growth trajectories by site, we also found site-specific differences by child age. Children in rural road and rural river communities experienced most of their postnatal growth faltering after six months. Six months is when dietary changes occur (e.g., the introduction of complementary foods) and nutritional needs increase, as well as changes in mobility (e.g. sitting, crawling), which can lead to increased environmental exposures such as mouthing objects, highest among 6–24-month-olds (21), and interactions with animals, shown in this population to increase with age (22). The growth spurts during the 7-12 month period are particularly important for attained height (23), and interventions during this period may be important to reverse the trends we found in the rural communities. This may include environmental specific interventions, including reducing exposure to animals and their feces, or nutrition-based interventions, such as supplementation (24). However, more research into context-specific community-level factors is needed, given the strong interactions between age and community type.

Exposures that were proxies of *Resources* and *Norms* had more observed associations with growth trajectories over the two years and across settings compared to downstream *Underlying Determinants*. Maternal height was most strongly associated with LGAZ (8), as has been found previously and would be expected due to maternal potential, but we also found associations between maternal height and postnatal growth. A large meta-analysis of birth cohorts found that postnatal growth trajectories stratified by maternal height progressed similarly and only 30% of the effect of maternal height on 24-month LAZ was mediated by birth characteristics (7), while another study modeling across several cohorts found 66% mediation with 18-month LAZ (8). We extended this by controlling for attained growth directly before each 3-month period and found associations between maternal height and change in LAZ postnatally, even through the 7–24-month period. Maternal height may be function of access to resources and generational effects in addition to genetic potential (25). Another study across 54 low- and middle-income countries found that maternal height was not only associated with low height-for-age but also with low weight-for-age and mortality (26). However, results from supplementation trials targeting only the prenatal period to improve child attained growth (beyond birth outcomes) show no effect at 6 months (27). Given these consistent associations, changing maternal height to improve the growth and health of subsequent generations will require a life course approach to women’s health (28), starting in early childhood, when response to interventions targeting attained height is strongest (2).

Other Enabling Determinants with associations with postnatal growth trajectories across ages and urbanicity included if the child was male and household wealth, both common risk factors for stunting across settings (3,7). Males may be at a higher predisposition for stunting due to biological processes(29), including increased nutritional needs, independent of social norms favoring male children (30). Household wealth can have trickle-down effects on food security to improve nutrition, household infrastructure to prevent infection from environmental exposures, and psychosocial effects on parental mental health which can perpetuate cycles of poverty. One variable commonly linked to growth faltering is low maternal education, which we did find some evidence of in our urban site, however, education was negatively associated with growth in our most remote site (rural river). In a meta-analysis of multiple studies, improving maternal education was found to reduce more stunting than improving maternal height, specifically for Latin America, however the Latin American sites in the meta-analysis were mostly urban and peri-urban (7). Potential explanations for this include opportunity costs of education versus developing skills needed for sustaining a lifestyle in more rural areas, differences in the meaning and application of skills gained in formal education, or a bias in selection towards individuals who were educated but returned to the communities. However, more research, including potentially qualitative work around the role of maternal education in these unique rural settings, is needed.

Season was associated with growth trajectories only in the 7–24-month period and only in rural communities, where we found diverging associations depending on whether villages had road access (worse growth in the dry season) or were connected only by river (worse growth in the rainy season). In this region of Ecuador, rural road communities are more dependent on seasonal labor. We found higher levels of food insecurity compared to the other sites, and thus they may be more susceptible to food shocks, which are more common in the dry season. In contrast, a higher proportion of households in rural river communities may utilize subsistence farming for food and may be less susceptible to shocks (31). Still, household infrastructure, including water, sanitation, and hygiene infrastructure, is poorer in rural river sites, which may mean that children are more susceptible to infectious diseases during the rainy season. A previous study in this cohort found the highest rates of infection among 6-month-olds in rural river communities (12), including enteroaggregative *E.coli*, *Shigella*, *Campylobacter (jejuni* and *coli)*, and *Giardia lamblia*, pathogens associated with growth faltering (32), and a systematic review found higher prevalence of these pathogens during higher temperatures (33). These results suggest that seasonal factors may interact with structural factors, in addition to infectious disease transmission pathways, to influence growth.

### Underlying Determinants

Animal exposure was consistently associated with change in growth across child ages and geographic locations. This is consistent with a Bangladesh study that found that free-roaming animals in the households contribute to fecal contamination of soil (34) and that soil is the major pathway by which children ingest fecal pathogens (35). Prior research analyzing domestic animals and child health has had mixed outcomes, with many showing improved child growth and women’s nutritional status in addition to increases in infectious disease outcomes and morbidity (36–38). However, many studies conflate exposure to animals and animal feces with animal ownership, which may be an inadequate proxy for exposure and can in fact have economic benefits (39). Our animal exposure variable was based on a validated tool and scoring system developed in this study location, the FECEZ Enteropathogens Index (22), which not only captures information on animal ownership, but also the presence of and maternal and child interaction with animals and their feces. The original analysis conducted for the creation of the scoring system found that maternal exposures to animals and their feces dominated the variability in the data and defined the child’s environment (22). We used the index as an analysis of overall exposure to animals, however, further investigation into specific animals and behaviors that contribute to growth faltering is warranted, especially in light of the nutritional and economic benefits of animal ownership. The association between animal exposure and growth faltering should be examined more closely; in addition to intestinal pathogens, systemic zoonotic infections that are difficult to diagnose may also play a role.

Nutrition-related factors were the only other *Underlying Determinants* that were associated with size at birth and postnatal growth. Reported moderate or severe food insecurity was associated with lower LGAZ, and has been associated with low birth weight and small-for-gestational age outcomes (40). In addition to birth outcomes, household food insecurity was associated with lower attained growth at 24 months in a meta-analysis (7). We did not find overall associations with postnatal growth, but found that food insecurity was associated with better trajectories in the rural river communities. In conjunction, exclusive breastfeeding for more than 50% of days in the first six months was only associated with growth in the rural communities, potentially showing that breastfeeding is more important in areas of more limited resources for early childhood nutrition. There was no difference in magnitude across developmental period; although weaning typically occurs between 6 and 12 months, immune-related factors passed through breastmilk may provide protection from early infection (41) or assist with earlier establishment of a healthy microbiome (42). Alternatively, caregivers that choose to exclusively breastfeed may also choose other factors that improve growth, or may be better nourished beginning *in utero*, as maternal nutrition is a factor for sustained breastfeeding.

### Limitations and strengths

Although ECoMiD was designed to capture a range of environmental conditions by enrolling across a gradient of urbanicity, strong correlations between environmental factors and community type and limited variability within communities precluded the inclusion of some environmental variables, such as improved water, or led to model non-convergence for others. Along the same lines, we were not powered to detect precise site-specific associations, and this, combined with higher measurement error in the rural sites and loss to follow-up, may have led to low power in our stratified analyses. Furthermore, there may be season-location-exposure interactions (e.g., stronger associations between food insecurity and growth during the dry season in rural-road settings), that we are not powered to detect. Lastly, our outcome of interest, ΔLAZ, is more sensitive to measurement error than attained LAZ, both from regression to the mean and joint measurement error. However, we attempted to mitigate this by adjusting for LAZ at the start of the period and using algorithms across longitudinal growth trajectories to detect anomalies (18,43).

A major strength of the ECoMiD cohort overall is the recruitment of individuals across a rural-urban gradient. Examining the influence of urbanicity on growth across a small geographic area provides the opportunity to assess community-level exposures in a region with similar political, cultural, and economic conditions. The longitudinal anthropometric measures every three months allowed us to isolate effects within specific, defined periods. Repeated measures of the outcome allowed us to control for time-invariant confounding at the child-level to gain insight on the timing of growth faltering. We also studied rich household and environmental data, including longitudinal data on food security, animal exposures, water quality, and daily data on breastfeeding versus other methods of feeding. We also assessed maternal and child hand contamination and drinking water quality at longitudinal timepoints with repeated measurements, a more robust measurements of these typically noisy measures. Additionally, the ability to examine birth length and gestational age obtained from birth cards provided a more rigorous assessment of associations with birth outcomes due to the substantial changes in anthropometry that occur in the first few weeks of life.

## CONCLUSIONS

Our results show that child growth is dynamic in the first 1000 days, multifactorial, and driven by geographic and structural factors. Despite children born similar heights, postnatal trajectories differed, suggesting there remain opportunities for interventions that support linear growth past six months of age. Our findings highlight that child growth trajectories after six months were highly influenced by their environments, including urbanicity, as well as maternal and demographic factors that may be proxies of the broader structural factors at play. Using the rural-urban gradient to evaluate smaller differences in factors under an otherwise similar context provided the opportunity to elucidate the relative importance of environmental conditions.

## Data Availability

All data produced in the present study are available upon reasonable request to the authors.

## Acknowledgements

We thank study participants and families for providing their time for this study. We thank ECoMiD data collection field staff for their hard work to collect the data for this study. We also thank Andrew Mertens for discussions of results and providing insight from his work.

