## Supplementary Materials for "Prenatal and postnatal environmental influences on child growth trajectories across a rural-urban gradient: an analysis of the ECoMiD longitudinal birth cohort"

Supplementary Information

*Supplementary methods*

**Contamination variables**

Handrinses were performed on mothers and children in 100 mL of distilled water in a 69-oz Whirlpak bag. Samples of drinking water (100 mL) were taken from the source and the same assays were performed as on handrinses after sodium thiosulfate added to neutralize any chlorine. For each survey timepoint, samples were collected on three non-consecutive days (all within one week of each other) and were measured for *E.coli* using Colilert and Petrifilm. Samples were categorized into 1-5 ordinal score for *E.coli* levels depending on Colilert and Petrifilm results (see table below). The score for all three measurements was averaged and re-binned into a semi-continuous score that approximates the log scale of concentration of *E.coli*. Semi-continuous scores were then averaged across timepoints within households.

| **Colilert result** | **Petrifilm result** | **Concentration of E.coli per 100ml** | **Ordinal score (assigned for each sample [3 per timepoint])** | **Semi-continuous score (assigned to average of 3 samples per timepoint)** |
|---|---|---|---|---|
| Absent | 0 | <1 CFU or MPN/100 ml | 1 | 1 |
| Present | 0 | 1-99 MPN/100 ml | 2 | 2.5 |
| Present | 1-9 CFU/1 ml | 100-999 CFU/100 ml | 3 | 4 |
| Present | 10-49 CFU/1 ml | 1000-4999 CFU/100ml | 4 | 5 |
| Present | ≥50 CFU/1ml | ≥5000 CFU/100ml | 5 | 6 |

Figure S1. Exposure variable collection timepoints and imputation strategy.

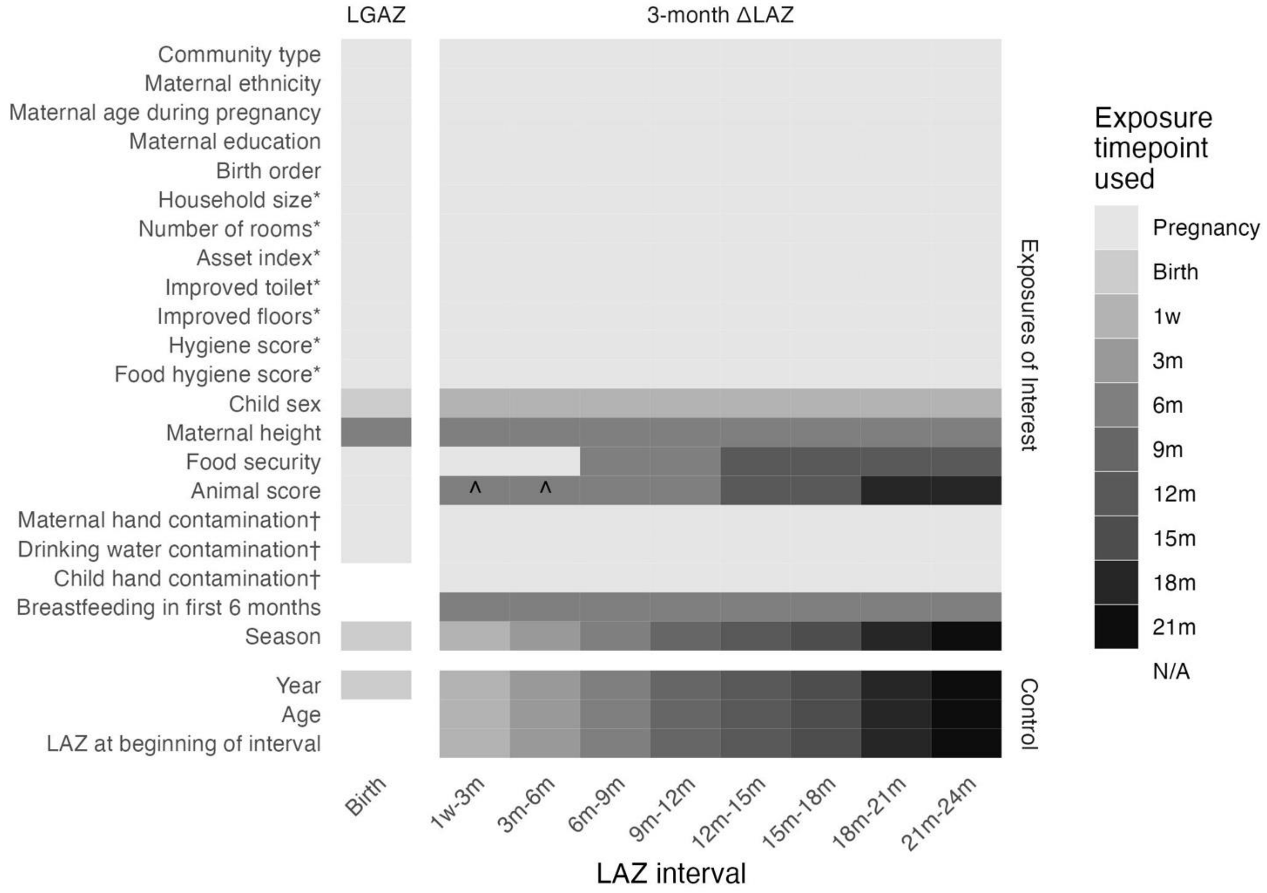


Notes: *Recollected if household moved but utilized pregnancy data only; ^Maternal exposure data; †Averaged over all visits

Table S1. Descriptive statistics of time-invariant exposure and outcome variables.

| | Overall<br>n = 401 | Urban<br>n = 102 | Intermediate<br>n = 115 | Rural – road<br>n = 142 | Rural – river<br>n = 42 |
|---|---|---|---|---|---|
| *Enabling Determinants – Norms* | | | | | |
| Child is male | 201 (51%) | 54 (54%) | 57 (50%) | 65 (46%) | 25 (60%) |
| Missing | 5 | 2 | 2 | 1 | 0 |
| Birth order | | | | | |
| 1 | 108 (27%) | 29 (28%) | 38 (33%) | 32 (23%) | 9 (21%) |
| 2 | 121 (30%) | 37 (36%) | 38 (33%) | 35 (25%) | 11 (26%) |
| 3+ | 172 (43%) | 36 (35%) | 39 (34%) | 75 (53%) | 22 (52%) |
| Maternal age | | | | | |
| Less than 22 yrs | 87 (22%) | 22 (22%) | 35 (30%) | 23 (16%) | 7 (17%) |
| 22-29 years | 210 (52%) | 52 (51%) | 55 (48%) | 85 (60%) | 18 (43%) |
| 30 years and more | 104 (26%) | 28 (27%) | 25 (22%) | 34 (24%) | 17 (40%) |
| Mom identifies as Afro-Ecuadorian | 290 (72%) | 51 (50%) | 72 (63%) | 126 (89%) | 41 (98%) |
| *Enabling Determinants – Resources* | | | | | |
| Household wealth quartile | | | | | |
| Q1 | 104 (26%) | 21 (21%) | 39 (34%) | 34 (24%) | 10 (24%) |
| Q2 | 97 (24%) | 22 (22%) | 26 (23%) | 34 (24%) | 15 (36%) |
| Q3 | 96 (24%) | 21 (21%) | 17 (15%) | 47 (33%) | 11 (26%) |
| Q4 | 104 (26%) | 38 (37%) | 33 (29%) | 27 (19%) | 6 (14%) |
| Maternal height (cm) | 160 (155, 164) | 160 (156, 164) | 158 (154, 164) | 160 (155, 166) | 158 (153, 162) |
| Missing | 37 | 18 | 10 | 8 | 1 |
| Maternal education | | | | | |
| Primary or less | 40 (10.0%) | 2 (2.0%) | 10 (8.7%) | 17 (12%) | 11 (26%) |
| Lower secondary | 60 (15%) | 3 (2.9%) | 15 (13%) | 27 (19%) | 15 (36%) |
| Upper secondary | 220 (55%) | 79 (77%) | 57 (50%) | 71 (50%) | 13 (31%) |
| Post-secondary or greater | 81 (20%) | 18 (18%) | 33 (29%) | 27 (19%) | 3 (7.1%) |
| *Underlying Determinants – Services* | | | | | |
| Number of rooms in household | 3 (2, 3) | 3 (2, 3) | 3 (2, 4) | 2 (2, 3) | 3 (2, 4.75) |
| Household size | 4 (3, 6) | 5 (3, 6) | 4 (3, 6) | 4 (3, 5) | 4 (3, 6) |
| Household has improved floor | 270 (67%) | 90 (88%) | 58 (50%) | 111 (78%) | 11 (26%) |
| Household has improved toilet | 375 (95%) | 101 (100%) | 108 (96%) | 137 (98%) | 29 (69%) |
| Missing | 5 | 1 | 2 | 2 | 0 |
| *Underlying Determinants – Practices* | | | | | |
| Hygiene score | 7 (5, 8) | 8 (5, 8) | 6 (5, 8) | 7 (5, 8) | 8 (8, 8) |
| Missing | 5 | 3 | 2 | 0 | 0 |
| Food covering score | 3 (3, 3) | 3 (3, 3) | 3 (2, 3) | 3 (3, 3) | 3 (3, 3) |
| Missing | 19 | 18 | 1 | 0 | 0 |
| Average maternal hand contamination | 2.0 (2.0, 2.5) | 2.5 (2.0, 2.5) | 2.5 (2.0, 2.5) | 2.0 (2.0, 2.5) | 1.5 (1.0, 1.5) |
| Missing | 46 | 8 | 9 | 21 | 8 |
| *Underlying Determinants – Food* | | | | | |
| Household is moderately/severely food insecure during pregnancy | 144 (36%) | 22 (22%) | 33 (29%) | 81 (57%) | 8 (19%) |
| Missing | 1 | 0 | 0 | 1 | 0 |
| Child was exclusively breastfed for >50% of days 0-6m | 211 (54%) | 58 (58%) | 54 (48%) | 65 (49%) | 34 (81%) |
| Missing | 13 | 2 | 2 | 9 | 0 |
| Average drinking water contamination | 2.5 (2.0, 2.5) | 2.5 (2.5, 2.5) | 2.5 (2.0, 2.5) | 2.0 (2.0, 2.5) | 1.5 (1.5, 2.0) |
| Missing | 46 | 8 | 9 | 21 | 8 |
| *Immediate Determinants – Care* | | | | | |
| Average child hand contamination | 2.5 (1.75, 2.5) | 2.5 (2.5, 2.5) | 2.5 (2.5, 2.5) | 2.5 (1.75, 2.5) | 1.0 (1.0, 1.0) |
| Missing | 77 | 24 | 16 | 28 | 9 |
| *Outcomes* | | | | | |
| Length-for-gestational-age Z-score, mean (SD) | 0.73 (1.27) | 0.61 (1.30) | 0.54 (1.27) | 0.92 (1.16) | 0.94 (1.45) |
| Missing | 22 | 3 | 9 | 2 | 8 |
| Average 3-month ΔLAZ, mean (SD) | -0.13 (0.81) | -0.09 (0.76) | -0.13 (0.79) | -0.13 (0.82) | -0.24 (-0.89) |

Note: Summary statistics presented are n (%) and median (IQR) unless noted otherwise

Table S2. Descriptive statistics of time-varying exposure and outcome variables (n = 401)

| | **Pregnancy** | **1w** | **3m** | **6m** | **9m** | **12m** | **15m** | **18m** | **21m** | **24m** |
|---|---|---|---|---|---|---|---|---|---|---|
| Household is moderately/severely food insecure | 144 (36%) | 144 (36%) | 144 (36%) | 129 (32%) | 129 (32%) | 133 (38%) | 133 (38%) | 132 (37%) | 132 (37%) | 126 (39%) |
| Missing | 1 | 1 | 1 | 0 | 0 | 47 | 47 | 47 | 47 | 81 |
| Animal exposure score | 1.61 (0.00, 3.51) | 0.48 (0.00, 2.29) | 0.48 (0.00, 2.29) | 0.48 (0.00, 2.29) | 0.48 (0.00, 2.29) | 1.35 (0.39, 3.02) | 1.35 (0.39, 3.02) | 1.67 (0.75, 2.75) | 1.67 (0.75, 2.75) | 1.58 (0.71, 2.45) |
| Missing | 0 | 34 | 34 | 34 | 34 | 46 | 46 | 70 | 70 | 89 |
| LAZ | - | 0.33 (-0.45, 1.04) | 0.21 (-0.62, 1.00) | 0.01 (-0.80, 0.75) | -0.36 (-1.10, 0.49) | -0.42 (-1.34, 0.40) | -0.67 (-1.60, 0.05) | -0.85 (-1.68, -0.07) | -1.06 (-1.73, -0.16) | -0.80 (-1.47, 0.03) |
| Missing | 401 | 12 | 42 | 55 | 65 | 69 | 75 | 100 | 96 | 98 |

Note: Summary statistics presented are n (%) and median (IQR) unless noted otherwise

Table S3. Unadjusted associations between community, demographic, and environmental exposures with child anthropometry at birth and child postnatal growth trajectories.

| | LGAZ | | | 3-month ΔLAZ | | |
|---|---|---|---|---|---|---|
| | n | MD | 95% CI | n | MD | 95% CI |
| *Enabling Determinants – Governance* | | | | | | |
| Community type | 378 | | | 2,477 | | |
| Urban | | — | — | | — | — |
| Intermediate | | -0.01 | -0.34, 0.33 | | -0.03 | -0.08, 0.02 |
| Rural – road | | 0.31 | 0.00, 0.62 | | -0.04 | -0.09, 0.01 |
| Rural – river | | 0.34 | -0.13, 0.81 | | -0.15 | -0.26, -0.05 |
| *Enabling Determinants – Norms* | | | | | | |
| Child is male | 378 | -0.11 | -0.35, 0.14 | 2,477 | 0.01 | -0.03, 0.05 |
| Birth order | 378 | | | 2,477 | | |
| 1 | | — | — | | — | — |
| 2 | | -0.23 | -0.55, 0.10 | | 0.05 | 0.00, 0.10 |
| 3+ | | -0.01 | -0.31, 0.30 | | 0.01 | -0.04, 0.06 |
| Maternal age during pregnancy | 378 | | | 2,477 | | |
| 30 years and more | | — | — | | — | — |
| Less than 22 yrs | | -0.04 | -0.40, 0.31 | | 0.01 | -0.04, 0.07 |
| 22-29 years | | 0.07 | -0.22, 0.37 | | 0.00 | -0.05, 0.05 |
| Mom identifies as Afro-Ecuadorian | 378 | 0.29 | 0.02, 0.56 | 2,477 | -0.01 | -0.05, 0.04 |
| *Enabling Determinants – Resources* | | | | | | |
| Household wealth quartile | 378 | | | 2,477 | | |
| Q1 | | — | — | | — | — |
| Q2 | | 0.03 | -0.32, 0.38 | | 0.03 | -0.03, 0.09 |
| Q3 | | -0.10 | -0.45, 0.24 | | 0.03 | -0.03, 0.09 |
| Q4 | | -0.13 | -0.47, 0.21 | | 0.08 | 0.03, 0.13 |
| Maternal height (per 5 cm) | 349 | 0.15 | 0.06, 0.24 | 2,458 | 0.01 | 0.00, 0.03 |
| Maternal education | 378 | | | 2,477 | | |
| Primary or less | | — | — | | — | — |
| Lower secondary | | -0.04 | -0.56, 0.47 | | -0.09 | -0.18, 0.00 |
| Upper secondary | | -0.01 | -0.44, 0.42 | | -0.04 | -0.11, 0.03 |
| Post-secondary or greater | | 0.01 | -0.48, 0.49 | | -0.05 | -0.12, 0.02 |
| Season | 378 | | | 2,477 | | |
| Late dry | | — | — | | — | — |
| Early rainy | | 0.15 | -0.25, 0.54 | | 0.04 | -0.06, 0.14 |
| Late rainy | | -0.10 | -0.46, 0.25 | | 0.07 | -0.03, 0.16 |
| Early dry | | -0.11 | -0.46, 0.23 | | 0.00 | -0.10, 0.09 |
| *Underlying Determinants – Services* | | | | | | |
| Number of rooms in household | 378 | 0.00 | -0.09, 0.09 | 2,477 | -0.01 | -0.03, 0.01 |
| Household size | 378 | 0.00 | -0.05, 0.05 | 2,477 | 0.00 | -0.01, 0.01 |
| Household has improved floor | 378 | 0.18 | -0.09, 0.44 | 2,477 | 0.01 | -0.04, 0.05 |
| Household has improved toilet | 374 | 0.28 | -0.31, 0.87 | 2,462 | -0.04 | -0.16, 0.08 |
| *Underlying Determinants – Practices* | | | | | | |
| Hygiene score | 374 | -0.01 | -0.08, 0.05 | 2,474 | 0.00 | -0.01, 0.01 |
| Food hygiene score | 361 | -0.07 | -0.23, 0.09 | 2,410 | 0.01 | -0.02, 0.03 |
| Animal exposure score | 378 | -0.04 | -0.16, 0.07 | 2,473 | -0.02 | -0.04, 0.00 |
| Average maternal hand contamination | 333 | 0.01 | -0.28, 0.29 | 2,194 | 0.05 | -0.01, 0.11 |
| *Underlying Determinants – Food* | | | | | | |
| Moderately/severely food insecure | 377 | -0.03 | -0.29, 0.23 | 2,473 | 0.05 | 0.00, 0.09 |
| Child was exclusively breastfed for >50% of days 0-6m | 370 | 0.02 | -0.23, 0.27 | 2,453 | 0.00 | -0.04, 0.04 |
| Average drinking water contamination | 333 | -0.23 | -0.52, 0.05 | 2,194 | 0.02 | -0.03, 0.07 |
| *Immediate Determinants – Care* | | | | | | |
| Average child hand contamination | | | | 2,178 | 0.07 | 0.01, 0.12 |

Note: MD = mean difference; CI = confidence interval; LGAZ = length-for-gestational-age Z-score; ΔLAZ = change in length-for-age Z-score.

Table S4. Effect modification of the association between community, demographic, and environmental exposures and birth anthropometry by community type.

| | | | Stratified Estimates | | | |
|---|---|---|---|---|---|---|
| Variable | Contrast | Global p-value | Urban | Intermediate | Rural - road | Rural - river |
| *Enabling Determinants – Norms* | | | | | | |
| Child is male | | 0.22 | -0.25 (-0.78, 0.28) | -0.57* (-1.06, -0.09) | 0.01 (-0.4, 0.43) | 0.24 (-0.62, 1.1) |
| Birth order | 2 | 0.12 | -0.41 (-1.08, 0.26) | 0.14 (-0.46, 0.74) | -0.57 (-1.2, 0.06) | 0.11 (-1.02, 1.24) |
| | 3+ | | -0.32 (-1.04, 0.4) | 0.27 (-0.38, 0.93) | -0.33 (-0.91, 0.26) | 1.18* (0.1, 2.26) |
| Maternal age during pregnancy | 22-29 years | 0.11 | 0.56 (-0.06, 1.19) | -0.1 (-0.72, 0.52) | 0.31 (-0.2, 0.83) | -0.68 (-1.63, 0.27) |
| | Less than 22 yrs | | 0.33 (-0.45, 1.11) | 0.07 (-0.66, 0.8) | 0.34 (-0.38, 1.07) | -1.4* (-2.59, -0.22) |
| Mom identifies as Afro-Ecuadorian | | 0.21 | -0.12 (-0.65, 0.41) | -0.16 (-0.66, 0.35) | 0.69* (0.02, 1.36) | 0.27 (-2.17, 2.71) |
| *Enabling Determinants – Resources* | | | | | | |
| Household wealth quartile | Q2 | 0.69 | -0.23 (-1.05, 0.58) | 0.11 (-0.56, 0.79) | 0.33 (-0.29, 0.94) | -0.03 (-1.09, 1.03) |
| | Q3 | | -0.24 (-1.05, 0.57) | 0.23 (-0.54, 1) | -0.21 (-0.78, 0.37) | -0.15 (-1.38, 1.07) |
| | Q4 | | -0.41 (-1.13, 0.31) | -0.34 (-0.96, 0.27) | 0.32 (-0.34, 0.98) | -0.44 (-1.78, 0.9) |
| Maternal height (per 5 cm) | | 0.28 | 0.03 (-0.22, 0.28) | 0.16 (-0.03, 0.35) | 0.18* (0.04, 0.33) | 0.39** (0.12, 0.66) |
| Maternal education | Lower secondary | 0.14 | 2.4* (0.05, 4.76) | -0.3 (-1.36, 0.76) | 0.21 (-0.56, 0.98) | -0.18 (-1.36, 1) |
| | Upper secondary | | 1.83* (0.12, 3.54) | -0.37 (-1.25, 0.51) | 0.53 (-0.16, 1.22) | -0.66 (-1.79, 0.48) |
| | Post-secondary or greater | | 1.54 (-0.29, 3.38) | -0.48 (-1.41, 0.46) | 0.73 (-0.04, 1.5) | -1.37 (-3.32, 0.59) |
| Season | Early rainy | 0.91 | -0.14 (-1.01, 0.74) | 0.17 (-0.69, 1.02) | 0.55 (-0.17, 1.26) | -0.19 (-1.35, 0.97) |
| | Late rainy | | -0.44 (-1.19, 0.31) | -0.27 (-1.07, 0.52) | 0.24 (-0.35, 0.83) | -0.97 (-2.58, 0.64) |
| | Early dry | | -0.37 (-1.07, 0.34) | -0.31 (-1.07, 0.45) | 0.15 (-0.44, 0.74) | -0.37 (-1.45, 0.71) |
| *Underlying Determinants – Services* | | | | | | |
| Number of rooms in household | | 0.035 | -0.1 (-0.39, 0.19) | -0.03 (-0.19, 0.14) | -0.19 (-0.41, 0.03) | 0.31* (0.04, 0.58) |
| Household size | | 0.36 | 0.07 (-0.04, 0.18) | -0.01 (-0.12, 0.09) | 0.04 (-0.07, 0.15) | 0.17 (-0.02, 0.36) |
| Household has improved floor | | 0.98 | 0.18 (-0.67, 1.02) | 0.14 (-0.38, 0.66) | 0.25 (-0.32, 0.82) | 0.33 (-0.65, 1.31) |
| *Underlying Determinants – Practices* | | | | | | |
| Hygiene score | | 0.87 | -0.06 (-0.27, 0.15) | -0.06 (-0.2, 0.07) | 0 (-0.13, 0.13) | 0.09 (-0.54, 0.72) |
| Food hygiene score | | 0.66 | -0.67 (-3.14, 1.8) | 0.08 (-0.22, 0.38) | -0.09 (-0.37, 0.2) | 0.27 (-0.44, 0.98) |
| Animal exposure score | | 0.82 | 0.07 (-0.27, 0.4) | -0.02 (-0.24, 0.21) | -0.11 (-0.33, 0.11) | -0.12 (-0.56, 0.32) |
| Average maternal hand contamination† | | 0.32 | 1.04* (0.06, 2.02) | 0.65 (-0.2, 1.51) | 0.22 (-0.4, 0.84) | -0.43 (-1.86, 1) |
| *Underlying Determinants – Food* | | | | | | |
| Moderately/severely food insecure | | 0.95 | -0.24 (-0.92, 0.44) | -0.37 (-0.92, 0.19) | -0.42 (-0.88, 0.04) | -0.12 (-1.27, 1.02) |
| Average drinking water contamination† | | 0.68 | -0.07 (-1.82, 1.68) | -0.03 (-0.71, 0.66) | -0.34 (-0.88, 0.21) | 0.49 (-0.83, 1.81) |

Note: *** p.value < 0.001; ** p.value < 0.01; * p.value < 0.05; aMD = adjusted mean difference; CI = confidence interval; LGAZ = length-for-gestational-age Z-score; ΔLAZ = change in length-for-age Z-score. Models are adjusted using the UNICEF categories: Model 1 includes Governance only, Model 2 includes all *Enabling Determinants*, Model 3 = all *Enabling Determinants* and *Underlying Determinants* except (†) variables, Model 4 = all *Enabling Determinants* and *Underlying Determinants, Model 5* = all *Enabling, Underlying,* and *Immediate Determinants.* All models are adjusted for year of measurement, and 3-month ΔLAZ models are adjusted for days between measurement, age at the beginning of the interval, and LAZ at the beginning of the interval.

Table S5. Effect modification of the association between community, demographic, and environmental exposures and postnatal growth trajectories by child age.

| | | | Stratified Estimates | |
|---|---|---|---|---|
| Variable | Contrast | Global p-value | 0-6 months | 7-24 months |
| *Enabling Determinants – Governance* | | | | |
| Community type | Intermediate | 0.00027 | -0.16 (-0.32, 0.01) | -0.13 (-0.27, 0.01) |
| | Rural - road | | -0.05 (-0.2, 0.11) | -0.16* (-0.3, -0.03) |
| | Rural - river | | -0.17 (-0.39, 0.06) | -0.64*** (-0.86, -0.42) |
| *Enabling Determinants – Norms* | | | | |
| Child is male | | 0.74 | -0.11 (-0.23, 0) | -0.13** (-0.23, -0.03) |
| Birth order | 2 | 0.84 | 0.05 (-0.12, 0.22) | 0.02 (-0.11, 0.16) |
| | 3+ | | -0.03 (-0.22, 0.16) | -0.07 (-0.23, 0.08) |
| Maternal age during pregnancy | 22-29 years | 0.042 | -0.11 (-0.25, 0.03) | -0.09 (-0.23, 0.04) |
| | Less than 22 yrs | | -0.12 (-0.31, 0.07) | 0.07 (-0.12, 0.25) |
| Mom identifies as Afro-Ecuadorian | | 0.0033 | 0.15 (0, 0.3) | -0.02 (-0.16, 0.11) |
| *Enabling Determinants – Resources* | | | | |
| Household wealth quartile | Q2 | 0.54 | 0.15 (-0.02, 0.33) | 0.09 (-0.05, 0.23) |
| | Q3 | | 0.08 (-0.08, 0.24) | 0.14 (-0.02, 0.3) |
| | Q4 | | 0.18* (0.02, 0.34) | 0.14* (0.01, 0.27) |
| Maternal height (per 5 cm) | | 0.59 | 0.09*** (0.04, 0.14) | 0.1*** (0.06, 0.14) |
| Maternal education | Lower secondary | 0.43 | -0.07 (-0.32, 0.19) | -0.13 (-0.35, 0.09) |
| | Upper secondary | | -0.13 (-0.35, 0.09) | -0.05 (-0.25, 0.15) |
| | Post-secondary or greater | | -0.24 (-0.48, 0) | -0.15 (-0.36, 0.06) |
| Season | Early rainy | 0.25 | 0 (-0.16, 0.16) | 0.13* (0.01, 0.25) |
| | Late rainy | | 0.07 (-0.1, 0.24) | 0.08 (-0.01, 0.18) |
| | Early dry | | -0.03 (-0.18, 0.11) | 0.1* (0.01, 0.2) |
| *Underlying Determinants – Services* | | | | |
| Number of rooms in household | | 0.33 | 0 (-0.05, 0.05) | -0.03 (-0.07, 0.02) |
| Household size | | 0.58 | 0 (-0.02, 0.03) | 0.01 (-0.02, 0.03) |
| Household has improved floor | | 0.72 | -0.02 (-0.16, 0.12) | 0 (-0.12, 0.13) |
| Household has improved toilet | | 0.29 | -0.19 (-0.49, 0.11) | -0.29* (-0.57, -0.01) |
| *Underlying Determinants – Practices* | | | | |
| Hygiene score | | 0.099 | 0.01 (-0.03, 0.04) | -0.02 (-0.05, 0.01) |
| Food hygiene score | | 0.67 | 0.02 (-0.06, 0.11) | 0.04 (-0.04, 0.12) |
| Animal exposure score | | 0.94 | -0.05 (-0.11, 0.01) | -0.05* (-0.09, -0.01) |
| Average maternal hand contamination† | | 0.0043 | -0.09 (-0.28, 0.1) | 0.14 (-0.02, 0.3) |
| *Underlying Determinants – Food* | | | | |
| Moderately/severely food insecure | | 0.58 | 0.05 (-0.06, 0.16) | 0.01 (-0.07, 0.1) |
| Child was exclusively breastfed for >50% of days 0-6m | | 0.8 | 0.1 (-0.02, 0.23) | 0.09 (-0.02, 0.19) |
| Average drinking water contamination† | | 0.098 | -0.04 (-0.19, 0.11) | 0.08 (-0.08, 0.23) |
| *Immediate Determinants – Care* | | | | |
| Average child hand contamination | | 0.0029 | -0.18* (-0.35, -0.02) | 0.02 (-0.13, 0.17) |

Note: *** p.value < 0.001; ** p.value < 0.01; * p.value < 0.05; aMD = adjusted mean difference; CI = confidence interval; LGAZ = length-for-gestational-age Z-score; ΔLAZ = change in length-for-age Z-score. Models are adjusted using the UNICEF categories: Model 1 includes Governance only, Model 2 includes all *Enabling Determinants*, Model 3 = all *Enabling Determinants* and *Underlying Determinants* except (†) variables, Model 4 = all *Enabling Determinants* and *Underlying Determinants, Model 5* = all *Enabling, Underlying,* and *Immediate Determinants.* All models are adjusted for year of measurement, and 3-month ΔLAZ models are adjusted for days between measurement, age at the beginning of the interval, and LAZ at the beginning of the interval.

Table S6. Effect modification of the association between community, demographic, and environmental exposures and postnatal growth trajectories by community type.

| Variable | Contrast | Global p-value | Stratified Estimates: Urban | Intermediate | Rural - road | Rural - river |
|---|---|---|---|---|---|---|
| *Enabling Determinants – Norms* | | | | | | |
| Child is male | | 0.076 | 0.01 (-0.2, 0.21) | -0.22** (-0.38, -0.06) | -0.19** (-0.33, -0.06) | 0.14 (-0.15, 0.43) |
| Birth order | 2 | 0.38 | -0.07 (-0.32, 0.18) | 0.13 (-0.06, 0.32) | -0.05 (-0.27, 0.16) | 0.15 (-0.39, 0.69) |
| | 3+ | | -0.32* (-0.62, -0.02) | 0.08 (-0.16, 0.32) | -0.07 (-0.27, 0.13) | 0.05 (-0.39, 0.5) |
| Maternal age during pregnancy | 22-29 years | 0.75 | 0.01 (-0.24, 0.26) | -0.24* (-0.48, 0) | -0.04 (-0.21, 0.13) | -0.2 (-0.53, 0.13) |
| | Less than 22 yrs | | 0.11 (-0.17, 0.4) | -0.13 (-0.41, 0.14) | 0.05 (-0.19, 0.29) | -0.11 (-0.67, 0.46) |
| Mom identifies as Afro-Ecuadorian | | 3.1e-35 | 0.01 (-0.2, 0.22) | -0.03 (-0.19, 0.14) | 0.37** (0.14, 0.59) | -1.3*** (-1.49, -1.11) |
| *Enabling Determinants – Resources* | | | | | | |
| Household wealth quartile | Q2 | 0.72 | -0.04 (-0.32, 0.25) | 0.28* (0.06, 0.51) | 0.05 (-0.16, 0.26) | 0.07 (-0.38, 0.52) |
| | Q3 | | 0.08 (-0.17, 0.33) | 0.23 (-0.06, 0.52) | 0.02 (-0.17, 0.21) | 0.05 (-0.45, 0.54) |
| | Q4 | | 0.15 (-0.09, 0.39) | 0.29** (0.09, 0.49) | 0.02 (-0.18, 0.22) | -0.08 (-0.67, 0.51) |
| Maternal height (per 5 cm) | | 0.86 | 0.11* (0.02, 0.21) | 0.07* (0.01, 0.14) | 0.09*** (0.04, 0.14) | 0.12* (0, 0.24) |
| Maternal education | Lower secondary | 0.015 | 0.58 (-0.06, 1.23) | -0.11 (-0.5, 0.27) | -0.03 (-0.31, 0.25) | -0.4* (-0.76, -0.05) |
| | Upper secondary | | 0.62* (0.07, 1.17) | 0.13 (-0.18, 0.44) | -0.06 (-0.33, 0.2) | -0.69*** (-1.09, -0.28) |
| | Post-secondary or greater | | 0.57 (-0.05, 1.19) | 0.02 (-0.31, 0.34) | -0.24 (-0.53, 0.04) | -0.45* (-0.84, -0.07) |
| Season | Early rainy | 0.00024 | 0.04 (-0.13, 0.22) | 0.01 (-0.15, 0.17) | 0.24** (0.08, 0.4) | -0.19 (-0.43, 0.04) |
| | Late rainy | | 0.05 (-0.12, 0.23) | -0.08 (-0.23, 0.07) | 0.32*** (0.19, 0.45) | -0.27* (-0.53, -0.01) |
| | Early dry | | -0.03 (-0.2, 0.14) | -0.07 (-0.21, 0.07) | 0.22** (0.09, 0.35) | -0.08 (-0.28, 0.12) |
| *Underlying Determinants – Services* | | | | | | |
| Number of rooms in household | | 0.4 | -0.07 (-0.15, 0.02) | 0.01 (-0.05, 0.06) | -0.01 (-0.09, 0.06) | -0.04 (-0.12, 0.03) |
| Household size | | 0.91 | 0 (-0.03, 0.04) | 0 (-0.03, 0.03) | 0 (-0.03, 0.04) | 0.02 (-0.03, 0.08) |
| Household has improved floor | | 0.98 | -0.03 (-0.22, 0.16) | 0 (-0.18, 0.18) | 0.01 (-0.19, 0.21) | -0.06 (-0.4, 0.28) |
| *Underlying Determinants – Practices* | | | | | | |
| Hygiene score | | 0.027 | -0.04 (-0.11, 0.03) | 0.01 (-0.04, 0.05) | 0 (-0.04, 0.04) | -0.27** (-0.45, -0.09) |
| Food hygiene score | | 8.5e-15 | 0.99*** (0.77, 1.22) | 0.02 (-0.06, 0.11) | 0 (-0.1, 0.1) | 0.19* (0.02, 0.35) |
| Animal exposure score | | 0.81 | -0.1 (-0.2, 0.01) | -0.05 (-0.11, 0.01) | -0.05 (-0.1, 0) | -0.01 (-0.18, 0.15) |
| Average maternal hand contamination† | | 0.42 | -0.01 (-0.39, 0.37) | 0.25 (-0.08, 0.58) | 0.01 (-0.19, 0.21) | -0.2 (-0.63, 0.24) |
| *Underlying Determinants – Food* | | | | | | |
| Moderately/severely food insecure | | 0.0043 | 0 (-0.13, 0.14) | -0.01 (-0.14, 0.12) | 0.01 (-0.11, 0.13) | 0.4*** (0.2, 0.6) |
| Child was exclusively breastfed for >50% of days 0-6m | | 0.12 | -0.07 (-0.29, 0.15) | 0.03 (-0.14, 0.2) | 0.2** (0.06, 0.35) | 0.3 (-0.07, 0.68) |
| Average drinking water contamination† | | 0.23 | 0.49 (-0.26, 1.25) | 0.13 (-0.14, 0.4) | -0.01 (-0.17, 0.15) | -0.34 (-0.82, 0.14) |
| *Immediate Determinants – Care* | | | | | | |
| Average child hand contamination | | 0.52 | -0.25 (-0.6, 0.1) | -0.14 (-0.48, 0.21) | -0.06 (-0.21, 0.09) | 0.19 (-0.27, 0.65) |

Note: *** p.value < 0.001; ** p.value < 0.01; * p.value < 0.05; aMD = adjusted mean difference; CI = confidence interval; LGAZ = length-for-gestational-age Z-score; ΔLAZ = change in length-for-age Z-score. . Models are adjusted using the UNICEF categories: Model 1 includes Governance only, Model 2 includes all *Enabling Determinants*, Model 3 = all *Enabling Determinants* and *Underlying Determinants* except (†) variables, Model 4 = all *Enabling Determinants* and *Underlying Determinants, Model 5* = all *Enabling, Underlying,* and *Immediate Determinants.* All models are adjusted for year of measurement, and 3-month ΔLAZ models are adjusted for days between measurement, age at the beginning of the interval, and LAZ at the beginning of the interval.
